# COVID-19 vaccine impact on rates of SARS-CoV-2 cases and post vaccination strain sequences among healthcare workers at an urban academic medical center: a prospective cohort study

**DOI:** 10.1101/2021.03.30.21254655

**Authors:** Tara C. Bouton, Sara Lodi, Jacquelyn Turcinovic, Sarah E. Weber, Emily Quinn, Cathy Korn, Jacqueline Steiner, Elissa M. Schechter-Perkins, Elizabeth Duffy, Elizabeth J. Ragan, Bradford P. Taylor, Beau Schaeffer, Nancy Miller, Ravin Davidoff, William P. Hanage, John Connor, Cassandra Pierre, Karen R. Jacobson

**Author notes:** Corresponding Author: Dr. Tara C Bouton, MD, MPH&TM, 801 Massachusetts Ave, 2^nd^ Fl, Boston, MA 02119.

## Abstract

**Background:** COVID-19 vaccine trials and post-implementation data suggest vaccination decreases SARS-CoV-2 infections. We examine COVID-19 vaccination’s impact on SARS-CoV-2 case rates and viral diversity among healthcare workers (HCW) during a high community prevalence period.

**Methods:** A prospective cohort study from Boston Medical Center (BMC)’s HCW vaccination program, where staff received two doses of BNT162b2 or mRNA-1273. We included PCR-confirmed SARS-CoV-2 cases among HCWs from December 09, 2020 to February 23, 2021. Weekly SARS-CoV-2 rates per 100,000 person-day overall and by time from first injection (1-14 and >14 days) were compared with surrounding community rates. Viral genomes were sequenced from SARS CoV-2 positive samples.

**Results:** SARS-CoV-2 cases occurred in 1.4% (96/7109) of HCWs given at least a first dose and 0.3% (17/5913) of HCWs given both vaccine doses. Adjusted SARS-CoV-2 infection rate ratios were 0.73 (95% CI 0.53-1.00) 1-14 days and 0.18 (0.10-0.32) >14 days from first dose. HCW SARS-CoV-2 cases >14 days from initial dose compared to within 14 days were more often older (46 versus 38 years, p=0.007), Latinx (10% versus 8%, p=0.03), and asymptomatic (48% versus 11%, p=0.0002). SARS-CoV-2 rates among HCWs fell below those of the surrounding community, with a 18% versus 11% weekly decrease respectively (p=0.14). Comparison of 48 SARS-CoV-2 genomes sequenced from post-first dose cases did not indicate selection pressure towards known spike-antibody escape mutations.

**Conclusions:** Our results indicate a positive impact of COVID-19 vaccines on SARS-CoV-2 case rates. Post-vaccination isolates did not show unusual genetic diversity or selection for mutations of concern.

**Main Point:** Cases of SARS-CoV-2 among health care workers dropped rapidly with COVID-19 vaccination. Sequencing 48 breakthrough infections (overwhelmingly in 14 days after 1st dose) showed no clear sign of any differences in spike protein compared with time-matched, unvaccinated control sequences.

## Introduction

On December 15, 2020, Boston Medical Center (BMC), an urban, safety-net hospital, started offering its 10,590 healthcare workers (HCWs) severe acute respiratory syndrome coronavirus 2 (SARS-CoV-2) vaccines BNT162b2 (Pfizer/BioNTech) and then mRNA-1273 (Moderna). The vaccination campaign coincided with Massachusetts’ second surge, which peaked at 6,000 new daily COVID-19 cases [1]. Vaccine was not available to anyone in the general population until February 1, 2021. This allowed examination of vaccine effectiveness during a period of higher prevalence and increased SARS-CoV-2 viral diversity than in the initial clinical trials [2, 3]. Concerns exist that newer SARS-CoV-2 variants have increased infectivity, modest decrease in neutralizing activity, and may impact vaccine effectiveness via escape from vaccine-induced immunity, specifically by mutations in the spike protein[4]. Assessment of viral genome sequencing of time-matched cases from vaccinated and unvaccinated individuals is needed to see whether such selection is evident.

Our aims were threefold: 1) to compare infection rates among HCWs who did and did not receive a SARS-CoV-2 vaccine, 2) to compare infection rates over time between HCWs and the surrounding community in the months following the BMC vaccination initiative and, 3) to compare genomic and spike protein mutations between cases detected post-vaccination and among unvaccinated cases.

## Methods

Boston University Medical Campus’ institutional review board approved the study. At BMC HCWs are screened daily for COVID-19 symptoms and tested if symptomatic. Asymptomatic testing is available to HCWs for workplace exposures, following out-of-state travel, and per request. Routine asymptomatic serial screening for SARS-CoV-2 infection was not performed during this period. All HCWs diagnosed with SARS-CoV-2 complete contact tracing and clinical questionnaires. As BMC was a BNT162b2 trial site, 66 HCWs had been vaccinated prior to the vaccine initiative and were included in analyses. The BMC COVID-19 vaccine initiative ultimately included all HCW, however rollout was staged, with patient-facing and employees caring for SARS-CoV-2 positive patients offered first.

We identified all HCWs with SARS-CoV-2 by reverse transcriptase-polymerase chain reaction (RT-PCR) between December 9, 2020 and February 23, 2021. HCW who received a vaccination following their positive SARS-CoV-2 RT-PCR were included in the unvaccinated group. We compared demographics and characteristics of cases detected post-vaccination by time since first dose to RT-PCR positive (1-14 days and >14 days from first dose to RT-PCR) using Pearson chi-squared, Fisher’s exact test, and Student t-tests. RT-PCR cycle threshold was dichotomized as above (negative) or below (positive) 24, the published cycle threshold above which SARS-CoV-2 virus has not been readily cultured, across 5 instruments (see Supplementary Table 1)[5–7]. We computed the crude weekly case rates by vaccination status at the time of the tests (unvaccinated, 1-14 days, and >14 days from first dose to RT-PCR) as the number of weekly cases divided by person-days of follow-up during that week (see Appendix for complete methodology). To control for confounding from community trends in infection, we adjusted the rates using direct standardization to the weekly rates in Middlesex, Norfolk, and Suffolk counties in Massachusetts [1]. We compared the weekly decline in rates after December 30, 2020 (14 days after the vaccine initiative started) for BMC HCWs and the community using a negative binomial regression model including an interaction term between week and group (BMC HCW versus community) and an offset for person-day at risk.

Residual isolates available from HCW SARS-CoV-2 cases tested at BMC were amplified using a modified ARTIC-primer based protocol and sequenced on an Illumina platform. Nucleotide substitutions, insertions, and deletions were identified with LoFreq [8] following alignment to the Wuhan Hu-1 reference sequence (NC_045512.2) [9]with Bowtie2[10]. We then restricted to cases from between January 1 and February 23, 2021 to time-match vaccinated and unvaccinated cases, which controlled for the expected accumulation of mutations due to the virus’s high mutation rate. Significance calculations for synonymous and nonsynonymous mutations between vaccination groups used a Wilcox non-parametric test. We further analyzed unique amino acid substitutions found in the spike protein of viruses isolated from cases >14 days after vaccination that were not found in unvaccinated cases, using initially an unmatched permutation analysis (see Supplementary Figure 1A) and then a date-matched permutation analysis (see Supplementary Figure 1B).

## Results

During the study period, 67% (7,109/10,590) of eligible HCWs were vaccinated with at least one dose. Post vaccination SARS-CoV-2 cases occurred in 96/7,109 (1.3%) HCWs who received at least one dose, 17/5,913 (0.3%) HCWs given both doses, and 329/3,481 (9.5%) unvaccinated HCWs. Seventy percent (67/96) of post vaccination SARS-CoV-2 cases occurred within 14 days of the initial dose (Figure 1). Comparison of HCW characteristics stratified by COVID-19 vaccination status at the time of SARS-CoV-2 positive RT-PCR are presented in Table 1, with post-initial vaccine dose cases more frequent among doctors and nurses compared to unvaccinated HCW cases (p<.01, Table 1). Among those who were vaccinated, HCWs diagnosed with SARS-CoV-2 >14 days after first dose were more often older (46 vs 38 years, p=0.007), Latinx (10% vs 8%, p=0.03), asymptomatic (48% vs 11%, p=0.0002), afebrile (100% vs 84%, p=0.02), and trended toward receipt of BNT162b2 (69% vs 48%, p=0.06), compared to HCWs diagnosed within 14 days of first dose (Table 2). There was no difference in RT-PCR cycle threshold. The adjusted rate ratios of SARS-CoV-2 infection were 0.73 (95% CI 0.53,1.00) and 0.18 (0.10,0.32) for 1-14 days and >14 days respectively from first dose compared to unvaccinated follow-up (Table 3).

**Figure 1:**
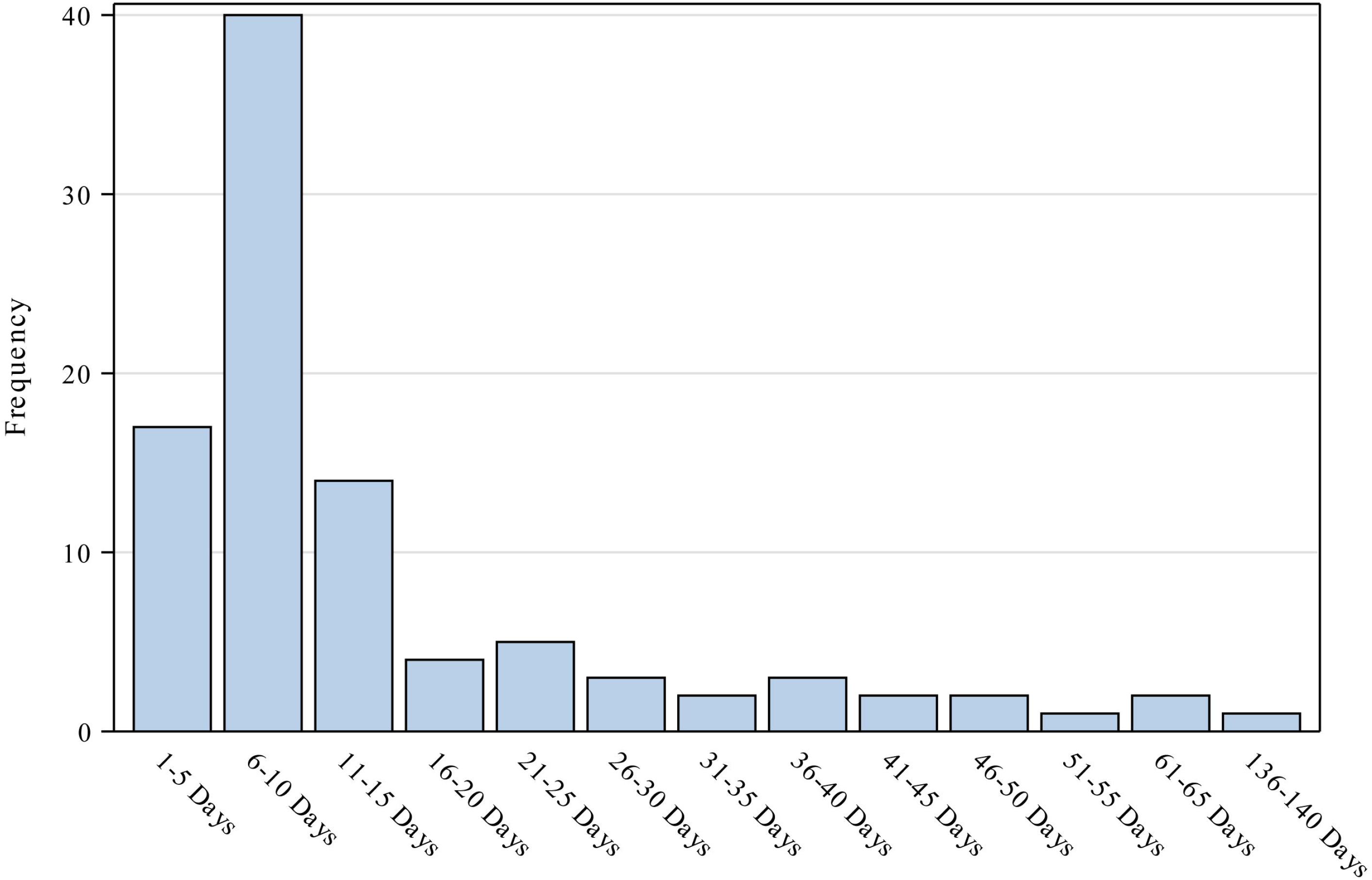
Time elapsed from first dose of SARS-CoV-2 vaccination to positive RT-PCR^a^ ^a^Abbreviations: SARS-CoV-2, Severe acute respiratory syndrome coronavirus 2; RT-PCR, reverse transcriptase-polymerase chain reaction

**Table 1:** Employee Characteristics Stratified by Vaccination Status at Time of Positive Test

| Characteristic, No. (%) | Total<br>(N=425) | Vaccination Status at Time of<br>Positive Test |  | P value<sup>a</sup> |
| --- | --- | --- | --- | --- |
|  |  | Unvaccinated<br>(N=329) | Post First<br>Vaccine Dose<br>(N=96) |  |
| **Age**, mean (SD), *years* (N=424) | 40 (13) | 39 (13) | 40 (13) | 0.53 |
| **Role** |  |  |  | <0.01 |
| Health care support worker | 56 (14) | 47 (15) | 9 (10) |  |
| Nurse | 95 (24) | 64 (21) | 31 (33) |  |
| Nurse Practitioner/Physician Assistant | 9 (2) | 4 (1) | 5 (5) |  |
| Administrative Staff | 42 (10) | 39 (13) | 3 (3) |  |
| Environmental Services | 7 (2) | 5 (2) | 2 (2) |  |
| Physician | 28 (7) | 15 (5) | 13 (14) |  |
| Medical technician | 22 (5) | 16 (5) | 6 (6) |  |
| OT, PT or speech therapist | 2 (1) | 1 (0.3) | 1 (1) |  |
| Pharmacy worker | 19 (5) | 14 (5) | 5 (5) |  |
| Respiratory Therapist | 4 (1) | 2 (1) | 2 (2) |  |
| Other | 120 (30) | 103 (33) | 17 (18) |  |
| **Test Reason** (N=375) |  |  |  | 0.48* |
| Community exposure | 130 (31) | 102 (31) | 28 (29) |  |
| Hospital exposure | 30 (7) | 21 (6) | 9 (9) |  |
| Unknown exposure | 215 (51) | 159 (48) | 56 (58) |  |
| **Asymptomatic** (N=375) | 88 (21) | 68 (21) | 20 (21) | 0.67* |
| **Fever** | 41 (10) | 30 (9) | 11 (12) | 0.49 |

Abbreviations: SD, standard deviation; OT, Occupational Therapist; PT, Physical Therapist; RT-PCR, reverse transcription-polymerase chain reaction
<sup>a</sup>Fisher's exact test, otherwise Pearson chi-squared or Student t-tests where appropriate

**Table 2:** Employee Characteristics Stratified by Vaccination-to-Diagnosis Timing

| Characteristic, No. (%) | Total<br>(N=96) | Days Between First Dose of<br>Vaccine and Positive Test |  | P value* |
| --- | --- | --- | --- | --- |
|  |  | 1-14 Days<br>(N=67) | 15+ Days<br>(N=29) |  |
| **Female** | 78 (82) | 54 (82) | 24 (83) | 1.00* |
| **Age**, mean (SD), years | 40 (13) | 38 (13) | 46 (13) | 0.007 |
| **Ethnicity** |  |  |  |  |
| Latinx | 8 (8) | 5 (8) | 3 (10) | 0.03* |
| **Race** |  |  |  | 0.12 |
| Asian/Asian Indian | 9 (10) | 6 (9) | 3 (10) |  |
| Black/African American | 21 (22) | 15 (23) | 6 (21) |  |
| Hispanic or Latino | 6 (6) | 5 (8) | 1 (3) |  |
| Native Hawaiian/Pacific Islander | 1 (1) | 0 (0) | 1 (3) |  |
| White | 50 (53) | 37 (57) | 13 (45) |  |
| Unknown/ Declined | 7 (7) | 2 (3) | 5 (17) |  |
| **Role** |  |  |  | 0.16 |
| Health care support worker | 9 (10) | 5 (8) | 4 (14) |  |
| Nurse | 31 (33) | 25 (38) | 6 (21) |  |
| Nurse Practitioner/Physician Assistant | 5 (5) | 4 (6) | 1 (4) |  |
| Administrative Staff | 3 (3) | 3 (5) | 0 (0) |  |
| Environmental Services | 2 (2) | 2 (3) | 0 (0) |  |
| Physician | 13 (14) | 6 (9) | 7 (25) |  |
| Medical technician | 6 (6) | 5 (8) | 1 (4) |  |
| OT, PT or speech therapist | 1 (1) | 1 (2) | 0 (0) |  |
| Pharmacy worker | 5 (5) | 3 (5) | 2 (7) |  |
| Respiratory Therapist | 2 (2) | 0 (0) | 2 (7) |  |
| Other | 17 (18) | 12 (18) | 5 (18) |  |
| **Type of Vaccine** |  |  |  | 0.06 |
| BNT162b2 | 52 (54) | 32 (48) | 20 (69) |  |
| mRNA-1273 | 44 (46) | 35 (52) | 9 (31) |  |
| **RT-PCR Cycle Threshold<sup>†</sup>** |  |  |  | 0.99 |
| >24 | 25 (26) | 18 (27) | 7 (24) |  |
| ≤24 | 39 (40) | 28 (42) | 11 (38) |  |
| Missing | 32 (33) | 21 (31) | 11 (38) |  |
| **Test Reason (N=93)** |  |  |  | 0.79* |
| Community exposure | 28 (30) | 19 (29) | 9 (34) |  |
| Hospital exposure | 9 (10) | 6 (9) | 3 (11) |  |
| Unknown exposure | 56 (60) | 41 (62) | 15 (56) |  |
| **Asymptomatic** | 20 (22) | 7 (11) | 13 (48) | 0.0002* |
| **Fever** | 11 (12) | 11 (16) | 0 (0) | 0.02 |

Abbreviations: SD, standard deviation; OT, Occupational Therapist; PT, Physical Therapist; RT-PCR, reverse transcription-polymerase chain reaction
\*Fisher's exact test, otherwise Pearson chi-squared or Student t-tests where appropriate
<sup>†</sup>24 was the lowest cycle threshold above which SARS-CoV-2 was unable to be cultured [5, 6, 7]

**Table 3:** SARS-CoV-2 rate reductions among BMC HCW by vaccination status^a^

| **SARS-CoV-2 Cases** | **Unvaccinated\*** | **Days Between First Dose of Vaccine and Positive Test** |  |
| --- | --- | --- | --- |
|  |  | **1-14 Days** | **15+ Days** |
| Number of cases | 329 | 67 | 29 |
| Person-days at risk | 406,387 | 96,041 | 251,790 |
| Crude rate per 100,000 person-day at risk (95% CI) | 80.96 (72.44, 90.20) | 69.76 (54.06, 88.60) | 11.52 (7.71, 16.54) |
| Adjusted rate per 100,000 person-day at risk (95% CI) | 71.64 (63.22, 80.07) | 50.02 (35.47, 64.57) | 12.69 (5.29, 20.09) |
| Adjusted rate ratio (95% CI) | Ref | 0.73 (0.53, 1.00) | 0.18 (0.10, 0.32) |

<sup>a</sup>See Appendix for full methodology

Following COVID-19 vaccine rollout to HCWs, case rates fell rapidly to below those of the community (Figure 2). Though not statistically significant, the observed weekly decrease in case rates was faster among BMC HCW than the community (HCW RR: 0.82; Community RR: 0.89; p=0.14).

**Figure 2:**
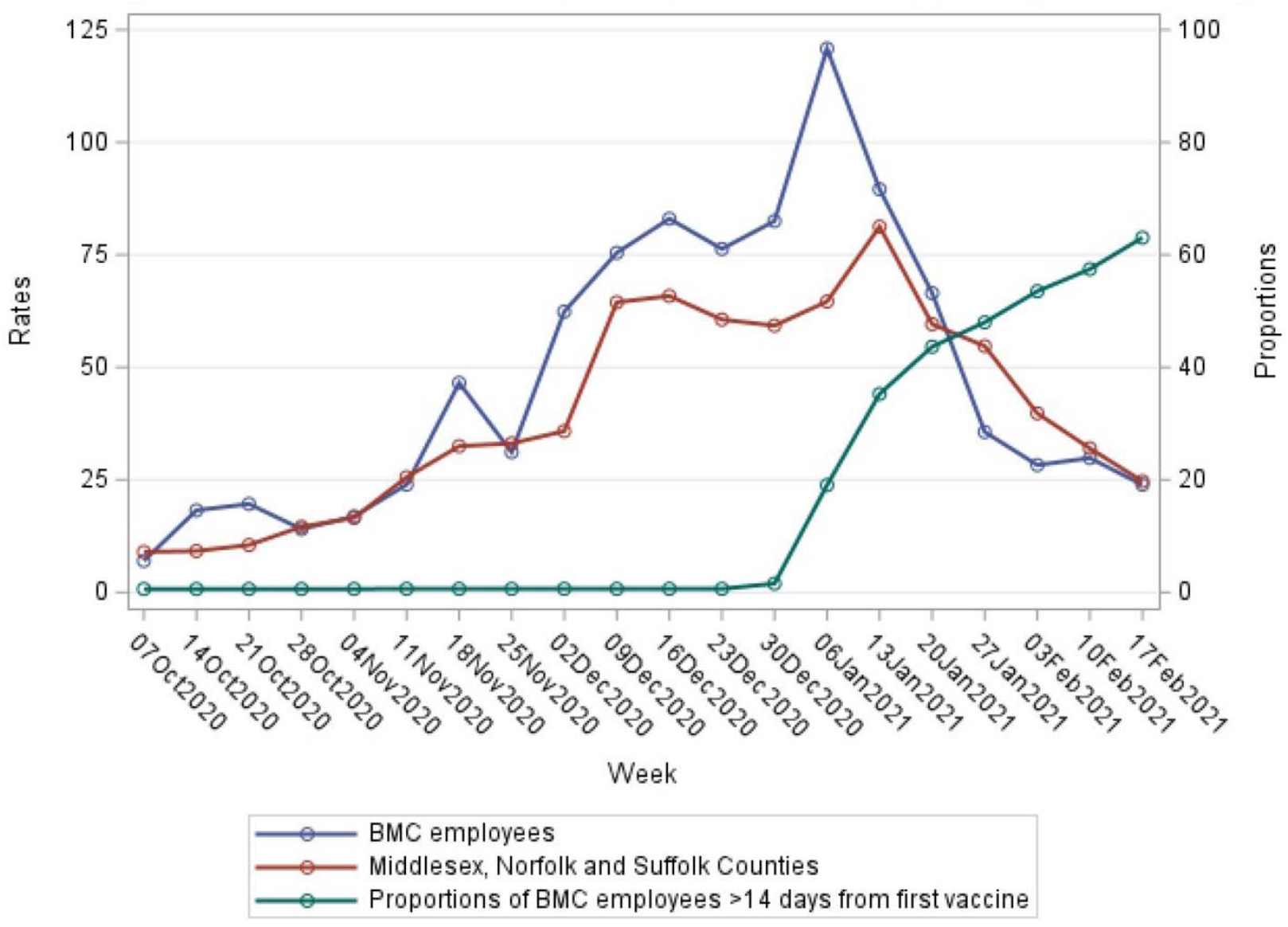
Weekly Rates of SARS-CoV-2 Cases per 100,000 Individuals and percent of vaccinated healthcare workers

We sequenced isolates from 50% (48/96) of SARS-CoV-2 cases diagnosed after at least the first vaccine dose. Sequences from 36 post-first vaccine SARS-CoV-2 cases and 51 unvaccinated cases were included in the time-matched analysis. Whole genome analysis identified 97 total nonsynonymous single nucleotide variants (SNVs) in the spike protein, with 64 and 68 SNVs among unvaccinated and post at least initial vaccine isolates (Figure 3A). Of these SNVs, 35 were seen in both the unvaccinated and vaccinated cases, consistent with these infections occurring in an environment where similar variants were circulating. To determine whether there was greater accumulation of SNVs in SARS-CoV-2 genomes from either unvaccinated or vaccinated groups, we plotted the distribution of nonsynonymous SNVs between genomes in each population (Figure 3B). This analysis showed that most SNVs were found in one single genome from an individual case, with little accumulation of multiple mutations in either population. The exception to this trend were three mutations associated with the D614G variant which appeared in all genomes analyzed.

**Figure 3:**
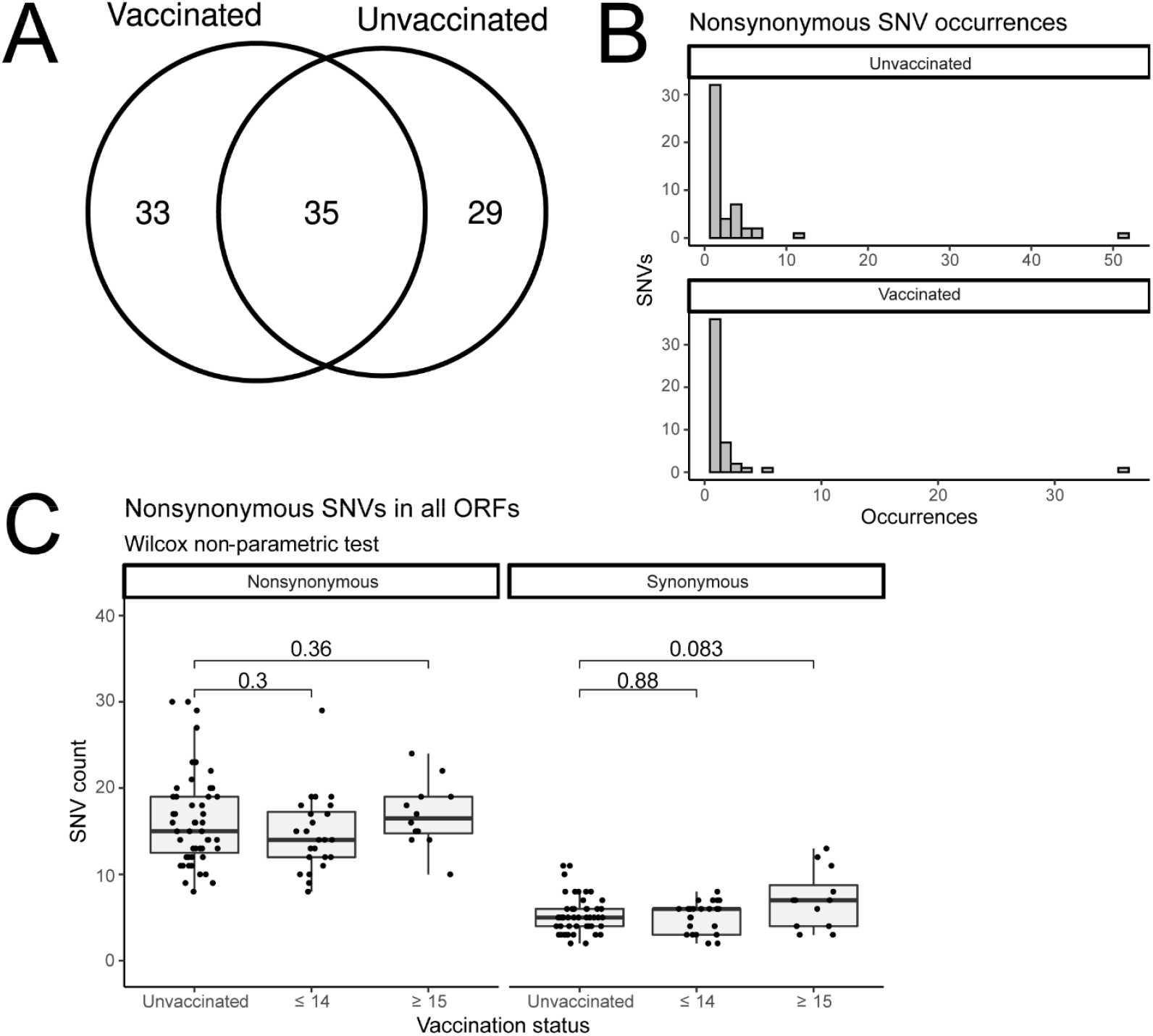
SARS-CoV-2 genome sequence variation among time-matched vaccinated and unvaccinated positive SARS-CoV-2 RT-PCR healthcare worker cases. A) Venn diagram illustrating the nonsynonymous SNVs in spike are distributed. A total of 97 SNVs were distributed over 87 time-matched sequenced genomes. 68 different SNVs were distributed over 36 genomes from individuals with a positive SARS-CoV-2 PCR following at least their first vaccination. 64 SNVs were distributed among 51 genomes from unvaccinated individuals. 35 SNVs were identified in genomes from both vaccinated and unvaccinated individuals. B) Distribution of spike SNVs between genomes in vaccinated and unvaccinated populations. C) Box-and-whiskers plots illustrating the number of nonsynonymous mutations found in infected HCW that were unvaccinated, infected less than 14 days after vaccination (≤ 14) or infected 15 or more days after vaccination as compared to the Wuhan-Hu-1 sequence. Significance calculations were calculated using a Wilcox non-parametric test.

Analysis of the amino acid changes seen in vaccinated cases identified none with the antibody-evading 484K mutation. There were two amino acid changes (T1117A and N121D) seen in two separate infections (see Supplementary Table 2). Phylogenetic analysis showed that in both cases these were two separate infections with the same closely related lineage, and not multiple separate acquisitions of the same mutation, which would suggest convergent evolution. While genome sequences from cases among vaccinees >14 days post first dose appeared more divergent from the original Wuhan-Hu-1 reference when compared with unvaccinated, this was no longer the case after controlling for the date of sampling; more recent samples are more divergent because more time has elapsed in which to accumulate mutations (Figure 3C). We further analyzed unique amino acid substitutions found in the spike protein of viruses isolated from cases >14 days after vaccination that were not found in unvaccinated cases. Although slightly fewer (p > 0.05) spike substitutions were observed in viruses isolated from cases >14 days after initial vaccine dose, adjusted for sampling date, we hypothesized that vaccine-induced selection pressure could be driving novel mutations. While an initial unmatched permutation analysis showed a significant difference (see Supplementary Figure 1A), a second date-matched permutation analysis was conducted and we concluded that the number of unique spike substitutions in viruses isolated from cases >14 days after the first dose was not significantly different than what would be expected by chance (see Supplementary Figure 1B).

## Conclusions

Our findings show SARS-CoV-2 vaccines decreased case rates at a time of high community prevalence and domestic circulation of concerning variants[11]. While post vaccination SARS-CoV-2 cases do occur, the majority are in the two weeks following the first dose. We found adjusted rate reduction of new cases in vaccinated HCWs in days 1-14 and >14 days post first dose versus unvaccinated of 27% and 82% respectively, similar to findings in the original clinical trials and more recently reported in Israel and other parts of the United States[12–17]. Vaccine protection starts to have a greater impact two weeks after the first vaccination dose. Individuals infected more than two weeks after first vaccination dose compared to before two weeks from first dose were older and reported fewer symptoms.

While we observed a decrease in rates among vaccinated versus unvaccinated individual HCWs, we did not observe a significant decrease in rates among HCWs versus the community in the months following the HCW vaccination initiative. Community rates likely surged in December due to holiday gatherings and travel, then trended down as individuals returned to more normal social distancing post the holiday season. Such behavior changes may have led to some of the HCW case rate drop as well, but the decline was steeper among HCWs likely reflecting vaccine impact. The HCW infection rates then flattened to falling community rates, even at a time when few members of the community were eligible for vaccine. HCWs at greatest risk for contracting SARS-CoV-2 infection outside of the workplace may have chosen not to receive vaccine, something we could not adjust for with our data, but could lead to the plateau in population-level effectiveness. Similarly, that we saw more post-vaccination cases among doctors and nurses likely reflects higher uptake of vaccine among HCW with those roles. We were unable to adjust for markers such as a social vulnerability index; however, disparities in vaccination uptake exist [18] and may lead to reduced vaccine effectiveness when compared with expectations from trial results. Even when access is not an obstacle, as in the case of HCWs, vaccine hesitancy and other barriers will need to be better understood and addressed to reach the levels of population immunity necessary to slow the spread of SARS-CoV-2 infection. In the meantime, mitigation measures such as masking, physical distancing, ventilation, and regular testing are still needed in hospital settings to protect both staff and patients. This is supported by recent modeling data suggesting that to avoid future outbreaks some mitigation measures will need to be maintained in the community at large even when high vaccine coverage is attained, as a result of the circulation of more transmissible variants[19].

Our data lacked follow-up from initial SARS-CoV-2 diagnosis, so initially asymptomatic cases may have progressed to symptomatic disease in all groups. Additionally, though HCWs diagnosed with SARS-CoV-2 more than 2 weeks from their initial dose were more likely to have received the BNT162b2 vaccine, this result likely reflects that BNT162b2 was given early when the force of infection was highest. Without routine asymptomatic screening for SARS-CoV-2 among BMC HCW during this time period, it is possible that unvaccinated BMC HCW were more likely to seek out asymptomatic testing than those who had received vaccine, leading to identification of more unvaccinated cases. An additional limitation of the study is that a larger sample size is likely needed to assess the statistical significance of the adjusted rate ratios of SARS-CoV-2 infection at 1-14 days.

There is intense interest in, and anxiety over, the potential of vaccination to select for mutants against which vaccines are less effective[20]. We found no early evidence of specific spike mutations or mutations associated with neutralizing vaccine escape, such as the E484K mutation. We observed overall genome divergence relative to the Wuhan-Hu-1 sequence as expected, with no evident increase of greater divergence in the Spike protein. Our study compared cases among vaccinees with those of contemporaneous unvaccinated individuals in the same population, which reflect the pool of virus to which vaccinated individuals were exposed, in order to avoid spurious associations between SNVs that happen to be locally common in different populations. It is a strength of this study that such time matched samples were available. Future studies should aim for similar design, as we had dramatically different findings when the unique spike protein substitution permutation analysis was conducted without date matching (see Supplementary Figure 1). We were also able to stratify participant samples by weeks from initial vaccine dose, when significant anti-spike antibody have been demonstrated to have developed[12, 21]. Not simply stratifying dichotomous vaccinated or unvaccinated samples, allowed observation of whether building immunity impacts the virus. Although we did not see clear trends, variation in the Spike sequence among vaccinated cases should be closely monitored by studies with larger sample size with careful consideration given to vaccine timing and comparison to concurrent unvaccinated controls.

Widespread vaccination does appear to impact case rates in real world campaigns. We must continue to improve vaccine coverage for all globally, educate about the need for vigilance in social distancing in the weeks following first dose, and carefully monitor for any ongoing viral evolution that impacts upon vaccine efficacy.

## Data Availability

A limited dataset is available from the authors by request.

## Acknowledgements

The authors would like to thank John Goldie for helping with the BMC vaccination data, Boston Medical Center for providing financial support for this study, and all the patients and staff involved in the BMC COVID-19 efforts this year. The biologic samples used for the analyses presented here were obtained from the BMC/BU COVID-19 Biorepository.

## Appendix

SARS-CoV-2 Rate Reductions Among BMC HCW by Vaccination Status: Full Methodology

Number of cases while unvaccinated, 1-14 days post vaccination and >14 days post vaccination were specified by week between December 9^th^ 2020 and February 24^th^ 2021. For each BMC employee, person-day of follow-up at risk while unvaccinated started on December 9^th^, 2020 and ended at the earliest of infection date, date of first vaccination shot or February 24^th^, 2021. Person-day follow-up at risk 1-14 days post vaccination started on the day after the first vaccination shot and ended at the earliest of infection date, the date corresponding to 14 after the first vaccination shot or end of the study. Person-day follow-up at risk >14 days post vaccination started 15 days after the first vaccination shot and ended at the earlier of infection date, or end of the study. Infection rates by vaccination status (unvaccinated, 1-14 post vaccination and >14 days post-vaccination) were expressed as number of cases per 100,000 person-day follow-up at risk overall and by week. Adjustment for temporal trends of infection in the community was performed by direct standardization to the weekly rates in 3 counties including and surrounding Boston where most BMC employees live. Directly standardized rates of the study population (unvaccinated BMC employees) are weighted averages of the weekly rates, where the weights are corresponding strata population size in the community. Weekly cases and estimated population in the 3 counties were obtained from the Massachusetts Department of Public Health publicly available dashboard and US Census Bureau, respectively and were used to compute the weekly infection rates in the community as number of cases per 100,000 person-days at risk.

**Supplementary Table 1:**
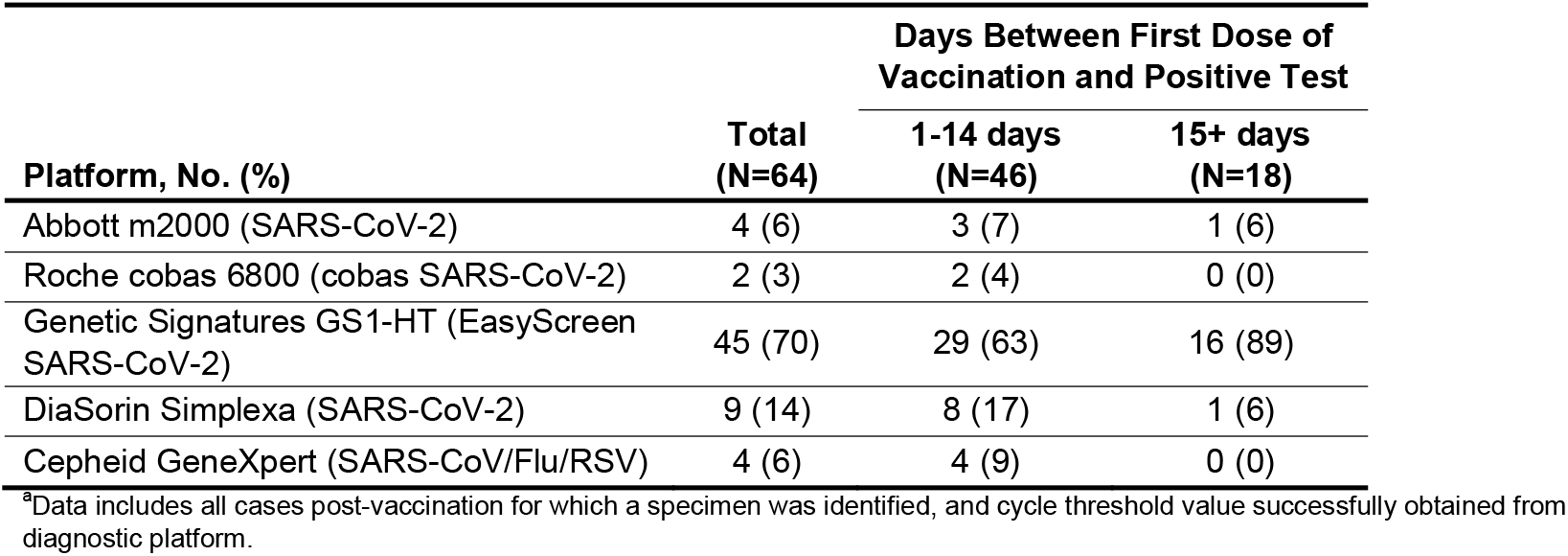
SARS-CoV-2 Platforms Used To Diagnose via RT-PCR, Cycle Threshold Values

**Supplementary Table 2.** Nonsynonymous mutations found in the Spike protein of vaccinated individuals who became infected (N=48)

| SNV | Count |
| --- | --- |
| N121D | 2 |
| T1117A | 2 |
| L8V | 1 |
| L18F | 1 |
| W152L | 1 |
| Q173K | 1 |
| Q271E | 1 |
| E309Q | 1 |
| P322S | 1 |
| T478K | 1 |
| N616D | 1 |
| A701V | 1 |
| T732A | 1 |
| G769V | 1 |
| E780Q | 1 |
| A845V | 1 |
| T859I | 1 |
| K1073R | 1 |
| H1101Y | 1 |
| I1232V | 1 |

**Supplementary Figure 1.**
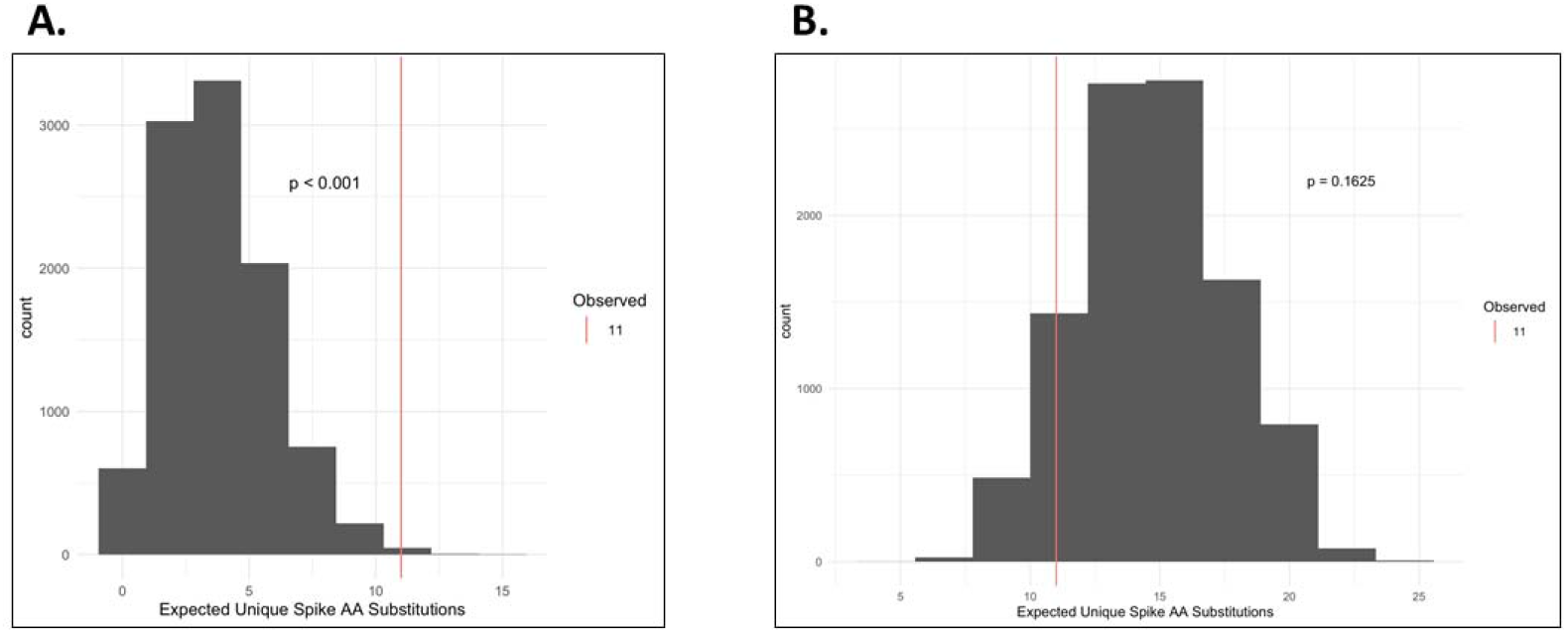
Permutation analysis of unique amino acid substitutions found in the spike protein of viruses isolated from cases >14 days after first vaccine dose Unmatched (A) and date-matched (B) permutation analysis of unique amino acid substitutions found in the spike protein of viruses isolated from cases diagnosed >14 days after their first vaccine dose that were not found in unvaccinated cases.

## References

1. Massachusetts Department of Public Health. COVID-19 Response Reporting. Mass.gov. 2021. https://www.mass.gov/info-details/covid-19-response-reporting%0D. Accessed 17 Feb 2021.

2. Wang Z, Schmidt F, Weisblum Y, Muecksch F, Barnes CO, Finkin S, et al. mRNA vaccine-elicited antibodies to SARS-CoV-2 and circulating variants. Nature. 2021. doi:10.1038/s41586-021-03324-6.

3. Greaney AJ, Loes AN, Crawford KHD, Starr TN, Malone KD, Chu HY, et al. Comprehensive mapping of mutations in the SARS-CoV-2 receptor-binding domain that affect recognition by polyclonal human plasma antibodies. Cell Host Microbe. 2021;S1931-3128:00082–2. doi:10.1016/j.chom.2021.02.003.

4. Abdool Karim SS, de Oliveira T. New SARS-CoV-2 Variants - Clinical, Public Health, and Vaccine Implications. N Engl J Med. 2021. doi:10.1056/NEJMc2100362.

5. Bullard J, Dust K, Funk D, Strong JE, Alexander D, Garnett L, et al. Predicting Infectious Severe Acute Respiratory Syndrome Coronavirus 2 From Diagnostic Samples. Clin Infect Dis. 2020;71:2663–6. doi:10.1093/cid/ciaa638.

6. Basile K, McPhie K, Carter I, Alderson S, Rahman H, Donovan L, et al. Cell-based culture of SARS-CoV-2 informs infectivity and safe de-isolation assessments during COVID-19. Clin Infect Dis. 2020. doi:10.1093/cid/ciaa1579.

7. Kim M-C, Cui C, Shin K-R, Bae J-Y, Kweon O-J, Lee M-K, et al. Duration of Culturable SARS-CoV-2 in Hospitalized Patients with Covid-19. N Engl J Med. 2021;384:671–3. doi:10.1056/NEJMc2027040.

8. Wilm A, Aw PPK, Bertrand D, Yeo GHT, Ong SH, Wong CH, et al. LoFreq: a sequence-quality aware, ultra-sensitive variant caller for uncovering cell-population heterogeneity from high-throughput sequencing datasets. Nucleic Acids Res. 2012;40:11189–201. doi:10.1093/nar/gks918.

9. Severe acute respiratory syndrome coronavirus 2 isolate Wuhan-Hu-1, complete genome. 2020. http://www.ncbi.nlm.nih.gov/nuccore/NC_045512.2. Accessed 28 Oct 2020.

10. Langmead B, Salzberg SL. Fast gapped-read alignment with Bowtie 2. Nat Methods. 2012;9:357–9. doi:10.1038/nmeth.1923.

11. United States Centers for Disease Control and Prevention. US COVID-19 Cases Caused by Variants. cdc.gov. 2021. https://www.cdc.gov/coronavirus/2019-ncov/transmission/variant-cases.html. Accessed 9 Mar 2021.

12. Polack FP, Thomas SJ, Kitchin N, Absalon J, Gurtman A, Lockhart S, et al. Safety and Efficacy of the BNT162b2 mRNA Covid-19 Vaccine. N Engl J Med. 2020;383:2603–15. doi:10.1056/NEJMoa2034577.

13. Baden LR, El Sahly HM, Essink B, Kotloff K, Frey S, Novak R, et al. Efficacy and Safety of the mRNA-1273 SARS-CoV-2 Vaccine. N Engl J Med. 2021;384:403–16. doi:10.1056/NEJMoa2035389.

14. Amit S, Regev-Yochay G, Afek A, Kreiss Y, Leshem E. Early rate reductions of SARS-CoV-2 infection and COVID-19 in BNT162b2 vaccine recipients. Lancet (London, England). 2021;397:875–7. doi:10.1016/S0140-6736(21)00448-7.

15. Keehner J, Horton LE, Pfeffer MA, Longhurst CA, Schooley RT, Currier JS, et al. SARS-CoV-2 Infection after Vaccination in Health Care Workers in California. N Engl J Med. 2021. doi:10.1056/NEJMc2101927.

16. Daniel W, Nivet M, Warner J, Podolsky DK. Early Evidence of the Effect of SARS-CoV-2 Vaccine at One Medical Center. N Engl J Med. 2021. doi:10.1056/NEJMc2102153.

17. Thompson MG, Burgess JL, Naleway AL, Tyner HL, Yoon SK, Meece J, et al. Interim Estimates of Vaccine Effectiveness of BNT162b2 and mRNA-1273 COVID-19 Vaccines in Preventing SARS-CoV-2 Infection Among Health Care Personnel, First Responders, and Other Essential and Frontline Workers - Eight U.S. Locations, December 2020-March. MMWR Morb Mortal Wkly Rep. 2021;70:495–500. doi:10.15585/mmwr.mm7013e3.

18. Burger AE, Reither EN, Mamelund S-E, Lim S. Black-white disparities in 2009 H1N1 vaccination among adults in the United States: A cautionary tale for the COVID-19 pandemic. Vaccine. 2021;39:943–51. doi:10.1016/j.vaccine.2020.12.069.

19. Moore S, Hill EM, Tildesley MJ, Dyson L, Keeling MJ. Vaccination and non-pharmaceutical interventions for COVID-19: a mathematical modelling study. Lancet Infect Dis. 2021. doi:10.1016/S1473-3099(21)00143-2.

20. Saad-Roy CM, Morris SE, Metcalf CJE, Mina MJ, Baker RE, Farrar J, et al. Epidemiological and evolutionary considerations of SARS-CoV-2 vaccine dosing regimes. Science. 2021. doi:10.1126/science.abg8663.

21. Krammer F, Srivastava K, Alshammary H, Amoako AA, Awawda MH, Beach KF, et al. Antibody Responses in Seropositive Persons after a Single Dose of SARS-CoV-2 mRNA Vaccine. N Engl J Med. 2021;384:1372–4. doi:10.1056/NEJMc2101667.

